# Predictive Modelling of Service Pathways to Admission in Psychiatric Residential Treatment Facilities

**DOI:** 10.1101/2022.07.22.22277941

**Authors:** Olga A. Vsevolozhskaya, Brian W. Turner, Stephen M. Shimshock, Kathi L.H. Harp, Xiaoran Tong, John S. Lyons

## Abstract

**Objective:** To develop and test predictive models of admissions to a psychiatric residential treatment facility (PRTF) in transitional age youth using routinely collected health insurance claims and enrollment data.

**Data Sources:** We used outpatient service and pharmaceutical claims from Medicaid beneficiaries aged 6-to 21-years old in Kentucky for the years 2010-2017.

**Study Design:** We assessed over 1,250 predictors (derived from Medicaid claims data) prior to the first PRTF admission. An ensemble machine learning (ML) algorithm based on logistic regression models fitted to a random subsample of the original data was used to predict pathways to the first PRTF admission. Discrimination performance of the ML ensemble was evaluated by comparing predictions to actual outcomes and calculating area under the curve (AUC), accuracy, sensitivity, and specificity. Additionally, a multivariate logistic regression model was fit to investigate the contribution of the continuity of mental health care after the initial PRTF admission on the risk of readmission.

**Data Collection/Extraction Methods:** We identified *N* = 519,011 unique children and youth with at least one outpatient service or pharmaceutical claim during our study period (January 1, 2010 through December 31, 2017).

**Principal Findings:** Fewer than 0.5% of children and youth in Kentucky had an episode of PRTF admission. Despite a very low prevalence of PRTF admission, classification accuracy of the ML ensemble for identifying PRTF youth achieved over 90% accuracy (AUC = 0.96). Factors associated with the initial PRTF admission were having been prescribed anti-psychotic and anti-manic medications, and receiving outpatient psychiatric care. Within six months after the initial PRTF discharge, there was a surprising drop in service utilization with a large proportion of youth not appearing to receive any follow-up care.

**Conclusions:** Despite the fact that admission into a PRTF was a relatively rare event, our findings suggest that it is a predictable event among youth with identified mental health conditions who are receiving care in the community.

**What is known on this topic:**

- After psychiatric hospitalization, PRTF treatment is the most expensive and restrictive intervention available to serve children and youth.
- Previous research examining predictors of PRTF entry using Medicaid reimbursement data showed that clinical factors were strong predictors of hospitalization.

**What this study adds:**

- We provide a comprehensive analysis of the factors beyond clinical diagnoses that lead to PRTF entry.
- We also seek to identify whether any specific patterns of service and/or pharmacy claims utilization are associated with reducing the likelihood of readmission.

## 1. INTRODUCTION

Centers for Medicare and Medicaid Services (CMS) define a psychiatric residential treatment facility (PRTF) as a non-hospital facility that provides inpatient services to Medicaid-eligible individuals under the age of 21 years.^1^ As one of only a handful of interventions that combine placement (i.e., a place to live) with treatment (i.e., a place to get help), PRTF interventions are most commonly utilized in situations where a placement is useful or required to potentiate treatment that cannot be managed with home- or community-based services. After psychiatric hospitalization, PRTF treatment is the most expensive and restrictive intervention available to serve children and youth, because these treatment settings provide specialized interventions, room and board, and 24 hour supervision.^2^ Historically, PRTFs were the only form of residential treatment fully funded by Medicaid. As such, residential treatment often represents a notable Medicaid expenditure within the broader child serving system. For example, a recent report found that roughly 8% of children and youth who receive mental health services come into contact with residential treatment, while, depending on the state, PRTF services account for 21-75% of Medicaid mental health expenditures.^2^ At the same time, other studies have found that the added cost and restrictions may not, on average, benefit children who are admitted relative to their peers treated in less-restrictive and less-costly family and community settings.^3,4^

Factors influencing concerns about the overuse of residential treatment go beyond its high cost. Issues like the danger posed to residents in PRTFs as a result of improper restraint and seclusion practices, and outdated approaches to treating children and youth with behavioral and emotional challenges have been attracting growing considerations.^5^ Under the 2019 Family First Legislation, a new emphasis is placed on raising children and youth in a family environment and guidelines are provided for the use of residential treatment in an effort to reduce unnecessary placements.^6^ Given these concerns, understanding factors that are associated with PRTF entry, thus elucidating pathways into residential treatment, is a priority consideration.

Previous research examining predictors of PRTF entry using Medicaid reimbursement data showed that clinical factors were strong predictors of hospitalization.^7^ Youths with mood, disruptive and psychotic disorder diagnoses, major depression, affective psychoses, and conduct disorders were commonly treated in PRTF.^2,7^ Other predictors of acute admission included prior hospitalization, receipt of two or more concurrent psychotropic medications, older age, and urban residence.^7^ A study examining predictors of PRTF entry among children involved with the child welfare system (CWS) found that trauma-related conditions, antipsychotic medication prescriptions, and entry into lower levels of out-of-home care were associated with PRTF entry, thus supporting the view that youth are admitted to PRTFs largely due to clinical need.^8^

The present study builds on the prior research on PRTF entry but also provides additional relevant insights in several important ways. First, we provide a comprehensive analysis of the factors beyond clinical diagnoses that lead to PRTF entry by exhaustively searching through over 1000 prescription drug medication claims and over 200 service utilizations from Medicaid reimbursement data prior to first PRTF placement. Second, since a subset of youth are admitted to multiple episodes of PRTF, we also seek to identify whether any specific patterns of service and/or pharmacy claims utilization are associated with reducing the likelihood of readmission. Our study goals are:

1. To build a machine learning model using predictive analytics algorithms to identify a parsimonious set of services and prescription medications that children and youth receive prior to admission to residential treatment.
2. To identify what (if any) services children and youth receive after exiting a residential treatment episode of care.

We hypothesize that the use of wraparound services or other intensive community services may impact the rate of admission into residential treatment. Further, we hypothesize that services received after exiting PRTF may be related to reduced likelihood of future readmissions.

## 2. METHODS

### 2.1 Data Source

Our person-level study used Kentucky Medicaid claims records from January 1, 2010 through December 31, 2017. These data contained information on all health services billed through Kentucky Medicaid insurance including eligibility file (provided Medicaid eligibility status, date of birth, gender, and race), paid pharmacy claims data file, and outpatient services file. The claims extract contained the quantity of drug dispensed, but no other information was provided on how it was prescribed. Therefore, we could not determine the total daily dose, frequency of administration, or duration of use, just the number of prescriptions that individual received. The outpatient services extract had a similar structure with only the number of services received. The University of Kentucky institutional review board approved this study.

### 2.2 Study Population

The initial data extract included claims for over *N* = 908,784 Medicaid recipients who were aged 0-31 years old as of December 31, 2017. Adults older than 21-year-old were excluded from the analyses, as well as children younger than six years old as no children under the age of six were admitted to a PRTF. A sample of children and youth with a paid pharmacy and outpatient services claims was selected for analyses of predictors to PRTF entry. This final sample included *N* = 519,011 unique children and youth.

### 2.3 Outcomes

We used the Medicaid claims data to identify children and youth admitted to a PRTF by using the billing provider type codes For the prediction modeling (i.e., the first study goal), we created a dummy variable identifying youth admitted to a PRTF (1 = admitted, 0 = not admitted or censored). For the analysis of children and youth after exiting a PRTF episode of care (i.e., the second study goal), we created a dummy variable identifying youth re-admitted to a PRTF (1 = re-admission, 0 = no re-admission or censored). Fewer than 0.5% of children and youth in the sample were admitted to a PRTF during the study period (*N* = 1,864), while 34% (*N* = 639) experienced at least one PRTF re-admission.

### 2.4 Predictors

From Medicaid pharmaceutical claims data, we identify 1,010 drug prescriptions that were reimbursed at least once by Medicaid for at least one out of *N* = 519,011 children and youth in our sample. From Medicaid outpatient services file, we identified 249 outpatient services that were reimbursed at least once by Medicaid for at least one of the sample participants. Then, we dichotomized the number of prescription drug reimbursements and the number of times a specific outpatient service was received as 1 = ever reimbursed and 0 = never claimed. The dichotomization was performed because the overwhelming majority of predictors had an extremely right-skewed distribution and this data transformation allowed us to ‘normalize’ them. For example, Abilify -- an antipsychotic drug, -- was never claimed by 512,284 youth in our sample, while at the same time its prescription was reimbursed 124 times for one individual. Despite common criticism of the dichotomization of a continuous measure prior to statistical analysis, there are situations in which dichotomized indicators perform as well as or better than the original continuous indictors and the extreme data skewness is one of these situations.^9^

Demographic covariates included age, gender, race, and ethnicity. Age was defined either as the age at the first PRTF entry or as the maximum age during the study period (for children and youth without PRTF entries).

### 2.4 Statistical Analyses

With *N* = 519,011 unique children and youth in our study, we had a sufficient sample size required to produce a stable solution when preforming a logistic regression analysis with ∼ 1,250 predictor variables. However, one challenge in using a logistic regression is the analysis of a binary outcome with a very low prevalence rate, because the analysis will often fail to produce meaningful results even if there are no numerical issues with the convergence of a likelihood function. For example, one can easily see how a logistic regression analysis of PRTF entries with < 0.5% prevalence rate may produce a stable but bogus solution by classifying all children to be non-PRTF cases, thus achieving over 99% accuracy (i.e., the percentage of children correctly classified by the model) with virtually zero sensitivity (i.e., the proportion of true positives).

#### 2.4.1 An Ensemble Machine Learning Approach

The ensemble models are the new generations of machine learning (ML) designed to provide higher classification accuracy than the conventional single model ML methods.^10,11^ Ensemble learning techniques have been shown to improve overall classification performance by combining an band of specialized weaker learners (i.e., models). These specialized learners are trained as separate classifiers using various subsets of the training data and then combined to form a weighted network of learners (i.e., the ensemble) that has a higher accuracy than any single learner.

In this study we used a logistic regression fitted to a random subsample of the original data as the base classifier, that is, the weak learner for the ensemble. Specifically, we randomly sampled 70% of all cases without replacement (i.e., 1,305 out of 1,864 PRTF admissions), and twice as many controls (i.e., 2,610 out of 517,147 non-PRTFs) resulting in a batch of *N* = 3,915 individuals with a 1:2 case/control ratio. Note that the batch sample size of *N* = 3,915 children and youth is not sufficient to produce a stable solution for a logistic regression with ∼ 1,250 predictors. To determine which predictor variables are relevant and should be included into the logistic regression model we computed their Renyi order-two entropy values.^12^ The entropy measures the homogeneity of utilization for each predictor for the subset of data; if a predictor’s entropy does not exceed a threshold, then the predictor is overly homogeneous for the subsample (i.e., too few claims for that medication or service among all 3,915 subsampled children), an entropy of zero means that only a single subject in the subsample utilized that medication or service, etc. For batch logistic regressions we included predictors with the top 10 percentile of entropy measures in their respective subsamples, resulting in 100 to 200 predictors for each of the base classifiers.

With this algorithm we created 1,000 batch models by fitting each base classifier on a random subsample of the original dataset and then combined the results of all the models to determine the final prediction.

#### 2.4.2 Prediction Performance Evaluation

To evaluate the performance of the ensemble ML model, we first estimated the probability of a PRTF entry for the remaining 30% testing data points in a batch. Since the original dataset is repetitively subsampled without replacement, certain subjects were selected many times to be in the testing set of different batches (e.g., all PRTF children), while others were selected a few times (e.g., some non-PRTF children). Therefore, each child had different number of predicted values for a PRTF entry. To make the final per-child prediction, we adopted a soft voting approach, where the prediction was made as a weighted sum with weights based on the base classifier accuracies. For example, assume a child was sub-sampled three times in testing data. Further assume that the three base logistic regression models resulted in estimated probabilities of 0.25, 0.40, and 0.27 for a PRTF entry. We first convert these probabilities to a binary 0/1 class as 0, 1, 0 (based on the 33% cut-off from the 1:2 case/control ration in a batch) and then combine them into the final prediction using the soft voting approach as *w*_l_ × 0 + *w*_2_ × 1 + *w*_3_ × 0, where *w*_l_, *w*_2_, and *w*_3_ are accuracies of logistic regressions that produced the estimated probabilities in the first place. In contrast, a hard-voting scheme would be just the majority vote (in the example above a vote of 0, since two out of three models predicted a zero).

Next, the performance of the ensemble ML was evaluated by tracking per-batch and the overall accuracy, sensitivity, specificity (i.e., the proportion of true negatives) and AUC-ROC curves. ROC is a probability curve that displays in a graphical way the trade-off between 1-sensitivity and specificity. AUC measures the ability of the classifier to distinguish between the 0 and 1 binary classes. A perfect classifier will have AUC of one, and worst performing classifier will have AUC of 0.5.

#### 2.4.3 Readmission Analysis

To investigate the contribution of the continuity of mental health care after the initial PRTF admission on the risk of readmission, we performed a multivariate logistic regression. The main predictors of interest were all pharmaceutical and service claims covered by Medicaid six months after the initial PRTF discharge with high entropy values. As we described above, predictors with low entropy were excluded because they represent rare events, i.e., claims that were made by very few subjects in the sample. The main outcome of interest was a binary flag for a PRTF readmission.

## 3. RESULTS

### 3.1 Sample Characteristics

Children and youth who were Medicaid beneficiaries during our study period were 51.7% female, 72.2% White and 11.9% African American, 95.6% non-Hispanic, with an average age of 12.6 years. The sample contained only a small portion of youth (*N* = 1,864, 0.36%) admitted to a PRTF during the study period. The median length of stay was 2.5 months, and 34% of children and youth (*N* = 639) experienced a subsequent PRTF readmission.

Table 1 reports univariate statistics as well as bivariate comparisons of children admitted to PRTFs during the study period with those who were not. Prior to the first PRTF admission, 3.6% did not claim any behavioral health services, 16.3% had claims for an emergency department visit, about 30% had claims for diagnostic psychiatric evaluations (i.e., [Psych Diagnostic Evaluation] 27.7% vs. 9.7%, *P* < 0.001, among children with no PRTF entries; [Psych Diag Eval W/Med Srvcs] 27.3% vs. 4.2%, *P* < 0.001; [Psy Dx Interview] 37.2% vs. 3.3%, *P* < 0.001), and over 50% had claims for psychotherapy services (i.e., psychotherapy with evaluation and management services [Psytx Office 20-30 Min] 68.2% vs. 10.2% in, *P* < 0.001; family psychotherapy with patient present [Family Psytx W/Pt 50 Min] 62.3% vs. 8.8%, *P* < 0.001; psychotherapy with explanations to the family [Consultation With Family] 70.2% vs. 11.7%, *P* < 0.001; psychotherapy focused on the monitoring and prescribing of psychopharmacologic agents [Medication Management] 55.3% vs 4.1%, *P* < 0.001). Regarding prescription drug claims among PRTF-admitted children, over 50% of them had at least one antipsychotic drug prescription (51% of children were prescribed risperidone and 33% Abilify (aripiprazole)), ∼35% had at least one prescription for an attention deficit hyperactivity disorder medication (ADHD) (37% clonidine, 35% guanfacine, 30% methylphenidate, 29% Vyvanse), and ∼30% had at least one antidepressant prescription (33% trazadone, 30% sertraline, 25% fluoxetine, 22% citalopram). Antibiotics and antihistamine medication prescriptions were the most utilized pharmaceutical claims among children and youth without a PRTF entry.

**Table 1.**
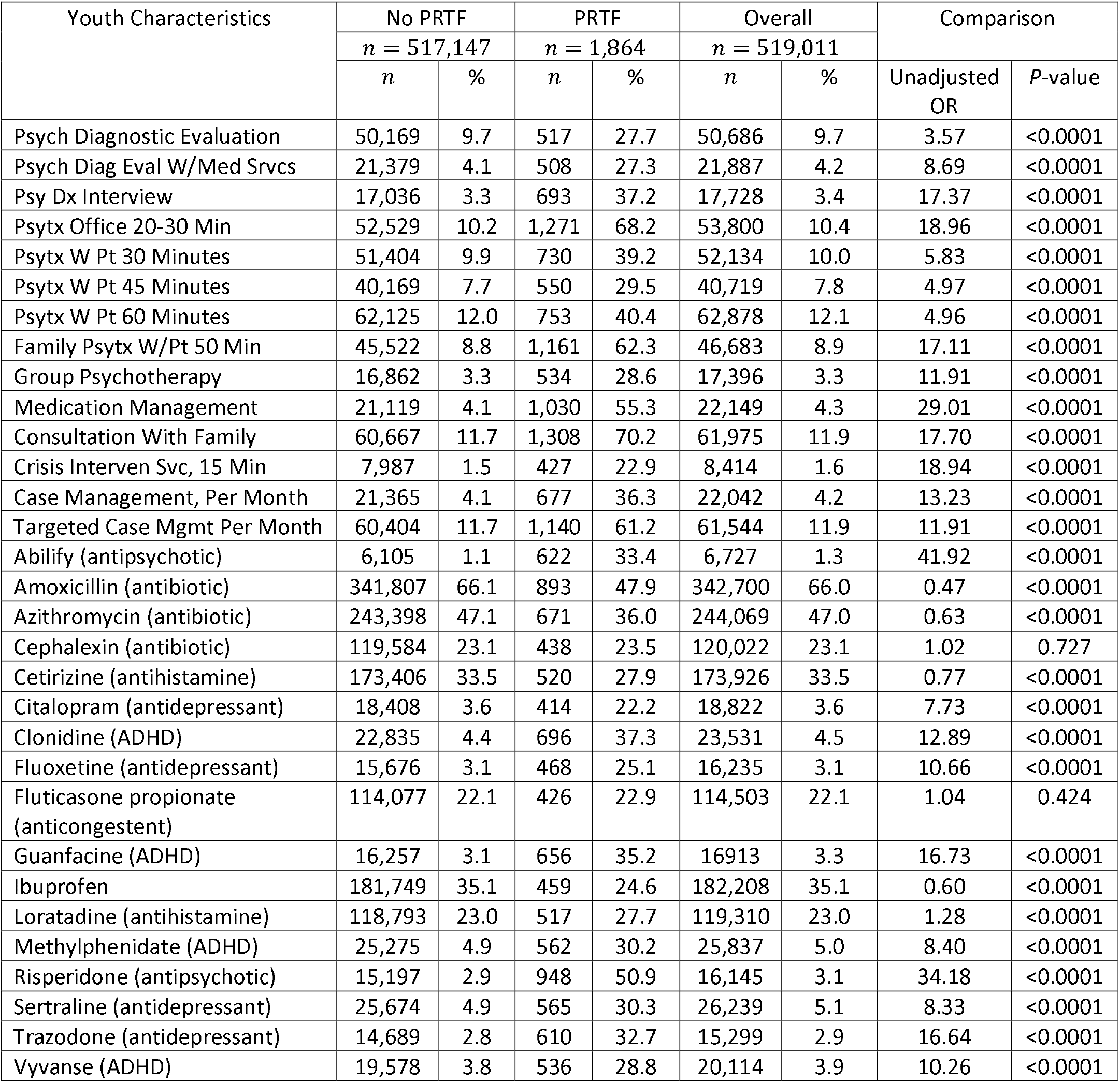
Description of the study sample with bivariate analyses comparing children with and without later PRTF admission by top behavioral health service and medication utilization.

| Youth Characteristics | No PRTF |  | PRTF |  | Overall |  | Comparison |  |
| --- | --- | --- | --- | --- | --- | --- | --- | --- |
|  | n = 517,147 |  | n = 1,864 |  | n = 519,011 |  | Unadjusted OR | P-value |
|  | n | % | n | % | n | % |  |  |
| Psych Diagnostic Evaluation | 50,169 | 9.7 | 517 | 27.7 | 50,686 | 9.7 | 3.57 | <0.0001 |
| Psych Diag Eval W/Med Svcs | 21,379 | 4.1 | 508 | 27.3 | 21,887 | 4.2 | 8.69 | <0.0001 |
| Psy Dx Interview | 17,036 | 3.3 | 693 | 37.2 | 17,728 | 3.4 | 17.37 | <0.0001 |
| Psytx Office 20-30 Min | 52,529 | 10.2 | 1,271 | 68.2 | 53,800 | 10.4 | 18.96 | <0.0001 |
| Psytx W Pt 30 Minutes | 51,404 | 9.9 | 730 | 39.2 | 52,134 | 10.0 | 5.83 | <0.0001 |
| Psytx W Pt 45 Minutes | 40,169 | 7.7 | 550 | 29.5 | 40,719 | 7.8 | 4.97 | <0.0001 |
| Psytx W Pt 60 Minutes | 62,125 | 12.0 | 753 | 40.4 | 62,878 | 12.1 | 4.96 | <0.0001 |
| Family Psytx W/Pt 50 Min | 45,522 | 8.8 | 1,161 | 62.3 | 46,683 | 8.9 | 17.11 | <0.0001 |
| Group Psychotherapy | 16,862 | 3.3 | 534 | 28.6 | 17,396 | 3.3 | 11.91 | <0.0001 |
| Medication Management | 21,119 | 4.1 | 1,030 | 55.3 | 22,149 | 4.3 | 29.01 | <0.0001 |
| Consultation With Family | 60,667 | 11.7 | 1,308 | 70.2 | 61,975 | 11.9 | 17.70 | <0.0001 |
| Crisis Interven Svc, 15 Min | 7,987 | 1.5 | 427 | 22.9 | 8,414 | 1.6 | 18.94 | <0.0001 |
| Case Management, Per Month | 21,365 | 4.1 | 677 | 36.3 | 22,042 | 4.2 | 13.23 | <0.0001 |
| Targeted Case Mgmt Per Month | 60,404 | 11.7 | 1,140 | 61.2 | 61,544 | 11.9 | 11.91 | <0.0001 |
| Abilify (antipsychotic) | 6,105 | 1.1 | 622 | 33.4 | 6,727 | 1.3 | 41.92 | <0.0001 |
| Amoxicillin (antibiotic) | 341,807 | 66.1 | 893 | 47.9 | 342,700 | 66.0 | 0.47 | <0.0001 |
| Azithromycin (antibiotic) | 243,398 | 47.1 | 671 | 36.0 | 244,069 | 47.0 | 0.63 | <0.0001 |
| Cephalexin (antibiotic) | 119,584 | 23.1 | 438 | 23.5 | 120,022 | 23.1 | 1.02 | 0.727 |
| Cetirizine (antihistamine) | 173,406 | 33.5 | 520 | 27.9 | 173,926 | 33.5 | 0.77 | <0.0001 |
| Citalopram (antidepressant) | 18,408 | 3.6 | 414 | 22.2 | 18,822 | 3.6 | 7.73 | <0.0001 |
| Clonidine (ADHD) | 22,835 | 4.4 | 696 | 37.3 | 23,531 | 4.5 | 12.89 | <0.0001 |
| Fluoxetine (antidepressant) | 15,676 | 3.1 | 468 | 25.1 | 16,235 | 3.1 | 10.66 | <0.0001 |
| Fluticasone propionate (anticongestent) | 114,077 | 22.1 | 426 | 22.9 | 114,503 | 22.1 | 1.04 | 0.424 |
| Guanfacine (ADHD) | 16,257 | 3.1 | 656 | 35.2 | 16,913 | 3.3 | 16.73 | <0.0001 |
| Ibuprofen | 181,749 | 35.1 | 459 | 24.6 | 182,208 | 35.1 | 0.60 | <0.0001 |
| Loratadine (antihistamine) | 118,793 | 23.0 | 517 | 27.7 | 119,310 | 23.0 | 1.28 | <0.0001 |
| Methylphenidate (ADHD) | 25,275 | 4.9 | 562 | 30.2 | 25,837 | 5.0 | 8.40 | <0.0001 |
| Risperidone (antipsychotic) | 15,197 | 2.9 | 948 | 50.9 | 16,145 | 3.1 | 34.18 | <0.0001 |
| Sertraline (antidepressant) | 25,674 | 4.9 | 565 | 30.3 | 26,239 | 5.1 | 8.33 | <0.0001 |
| Trazodone (antidepressant) | 14,689 | 2.8 | 610 | 32.7 | 15,299 | 2.9 | 16.64 | <0.0001 |
| Vyvanse (ADHD) | 19,578 | 3.8 | 536 | 28.8 | 20,114 | 3.9 | 10.26 | <0.0001 |

Overall, the bivariate results in Table 1 show that youth admitted to PRTFs were significantly more likely, prior to admission, to undergo a diagnostic psychiatric evaluation, to have some psychotherapy service utilization, and to have at least one psychotropic drug prescription.

### 3.2 Predictive Model Performance

Across 1,000 batches, the median base-model accuracy (i.e., the rate of correct predictions) was 91.7% with interquartile range (IQR), 91.0% - 92.5%; the median sensitivity (i.e., the true positive rate) was 88.7%, IQR 87.4% - 90.4%; the median specificity (i.e., the true negative rate) 93.3%, IQR 92.4% - 94.1%. Figure 1 shows ROC curves (i.e., sensitivity against 1-specificity) for a random realization of 100 base models. The overall ensemble, based on a weighted network of learners, attained 92.6% accuracy, 93.2% sensitivity, 92.6% specificity, and AUC value of 0.959 (highlighted in red in Fig. 1).

**Figure 1.**
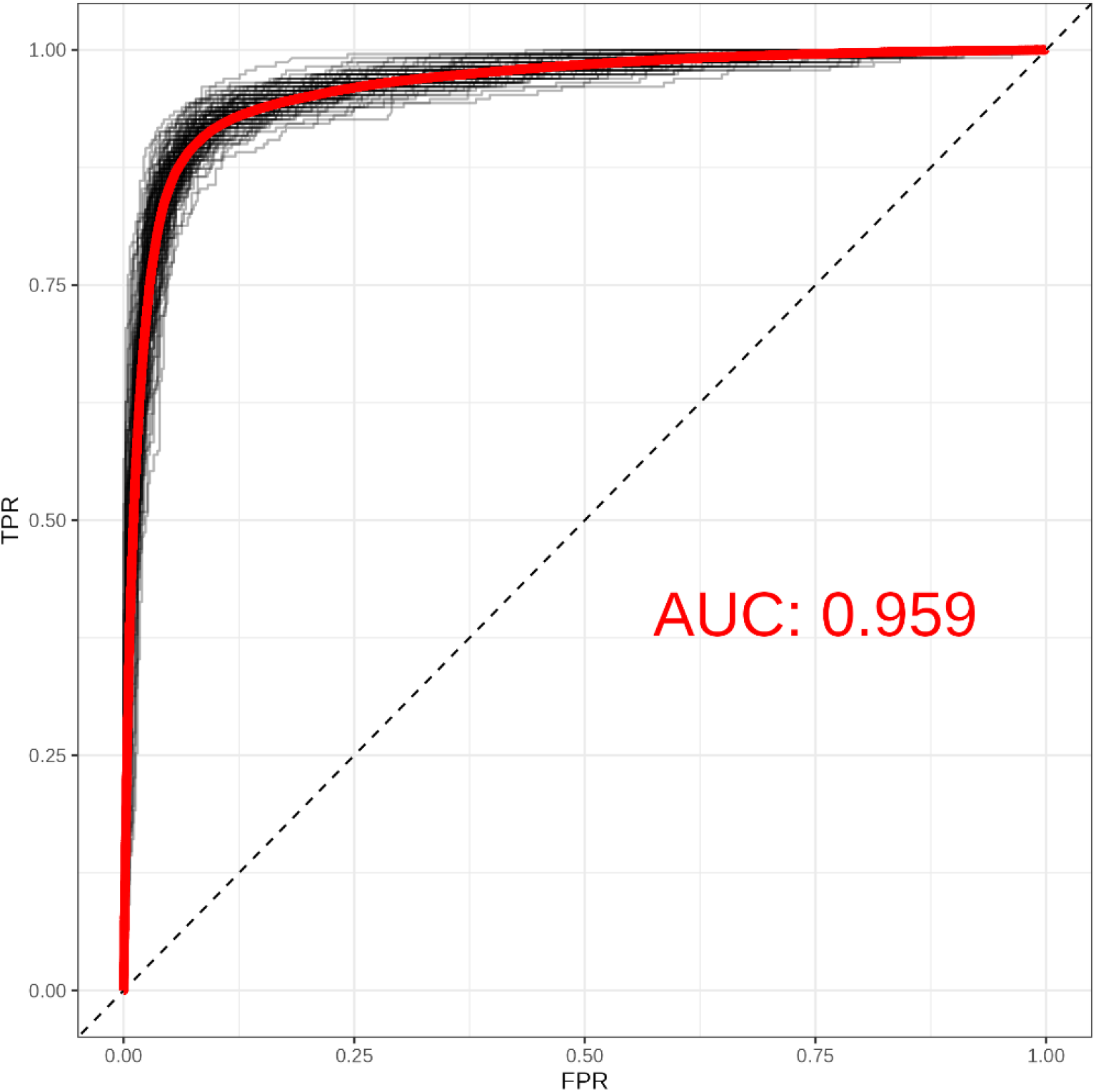
ROC curves (graph of sensitivity against 1-specificity) for a random sample of 100 base logistic models in the testing samples. The overall ROC-AUC results of per-child predictions for the entire ensembled are highlighted in red color.

#### 3.2.1 Factors Associated with a PRTF Entry

Table 2 summarizes top predictors across 1000 batches in terms of their median (IQR) adjusted odds ratios (aOR) stratified by age group. In general, service utilization and pharmaceutical claims demonstrated much stronger associations with a PRTF entry than did demographic characteristics with the exception of age. Age was a significant risk factor of PRTF entry among children ages 6- to 15-years old (6-12 years old, aOR = 1.77 IQR 1.61 – 1.98; 13-15 years old, aOR = 1.54 IQR 1.33 – 1.75), but a protective factor among older youth 16- to 21-years old (aOR = 0.16 IQR 0.12 – 0.21).

**Table 2.** Selected variables by predictive models and adjusted odds ratio of risk of PRTF entry.

| Youth Characteristics | Median Adjusted OR | IQR Adjusted OR |
| --- | --- | --- |
| **Children Ages 6- to 12-Years-Old** |  |  |
| Abilify (antipsychotic) | 8.39 | 3.81 – 19.05 |
| Age | 1.77 | 1.61 – 1.98 |
| Antipyrinebenzocaine (pain reliver) | 6.48 | 3.19 – 13.01 |
| Clonidine (ADHD) | 7.35 | 4.30 – 13.34 |
| Desmopressin acetate (diabetic) | 8.55 | 3.48 – 26.28 |
| Guanfacine (ADHD) | 15.98 | 8.71 – 33.93 |
| Oxcarbazepine (antiepileptic) | 12.31 | 4.97 – 34.95 |
| Risperidone (antipsychotic) | 21.41 | 12.46 – 43.39 |
| Psytx w pt 30 minutes | 28.16 | 9.45 – 105 |
| Psytx w pt 45 minutes | 4.61 | 3.06 – 7.95 |
| Analgesia | 4.76 | 2.85 – 7.87 |
| Mh health assess by non-md | 7.39 | 4.53 – 12.26 |
| Crisis intervention mental h | 7.95 | 5.55 – 11.62 |
| Amoxicillin (antibiotic) | 0.31 | 0.22 – 0.44 |
| Azithromycin (antibiotic) | 0.31 | 0.19 – 0.48 |
| Benzonatate (cough suppressant) | 0.02 | 0.01 – 0.03 |
| Hydrocodoneacetaminophen (opioid pain reliver) | 0.06 | 0.02 – 0.13 |
| Ibuprofen | 0.26 | 0.16 – 0.43 |
| Methylprednisolone (cortisone-like medicine or steroid) | 0.03 | 0.01 – 0.09 |
| Naproxen (nonsteroidal anti-inflammatory drug) | 0.01 | 0.00 – 0.02 |
| Ranitidine (antihistamine) | 0.14 | 0.06 – 0.26 |
| **Children Ages 13- to 15-Years Old** |  |  |
| Abilify (antipsychotic) | 5.73 | 3.78 – 8.84 |
| Age | 1.54 | 1.33 – 1.75 |
| Citalopram (antidepressant) | 4.18 | 2.81 – 6.40 |
| Clonidine (ADHD) | 3.22 | 2.30 – 4.46 |
| Fluoxetine (antidepressant) | 7.07 | 4.75 – 10.66 |
| Drug screen singl | 7.91 | 4.97 – 13.43 |
| Oxcarbazepine (antiepileptic) | 3.56 | 2.24 – 6.05 |
| Risperidone (antipsychotic) | 5.29 | 3.82 – 7.41 |
| Psytx office 20-30 min | 14.53 | 8.38 – 28.02 |
| Sertraline (antidepressant) | 4.50 | 3.29 – 6.42 |
| Psytx w pt 30 minutes | 3.01 | 2.41 – 3.77 |
| Trazodone (antidepressant) | 3.59 | 2.65 – 5.12 |
| Consultation with family | 2.57 | 1.90 – 3.47 |
| Analgesia | 2.73 | 2.07 – 3.64 |
| Drug screen multiple class | 2.94 | 2.19 – 3.84 |
| Crisis interven svc, 15 min | 2.73 | 2.07 – 3.85 |
| Crisis intervention mental h | 4.58 | 3.75 – 5.59 |
| Ciprofloxacin (antibiotics) | 0.13 | 0.39 – 0.57 |
| Hydrocodoneacetaminophen (opioid pain reliver) | 0.32 | 0.22 – 0.45 |
| IBU | 0.23 | 0.13 – 0.38 |
| Ibuprofen | 0.34 | 0.26 – 0.43 |
| Metronidazole (antibiotic) | 0.18 | 0.11 – 0.31 |
| Nystatin (antifungal) | 0.24 | 0.17 – 0.34 |
| Amoxicillin (antibiotic) | 0.48 | 1.40 – 1.74 |
| **Children Over 16-Year-Old** |  |  |
| Abilify (antipsychotic) | 31.88 | 13.31 – 123.01 |
| Fluoxetine (antidepressant) | 14.19 | 6.56 – 36.01 |
| Psych diag eval w/med srvc | 14.83 | 5.27 – 55.18 |
| Lamotrigine (anticonvulsant) | 9.81 | 3.70 – 30.24 |
| Quetiapine fumarate (antipsychotic) | 17.23 | 6.47 – 67.31 |
| Risperidone (antipsychotic) | 9.53 | 4.38 – 24.23 |
| Psytx off 20-30 min w/e&m | 17.59 | 4.52 – 83.16 |
| Sertraline (antidepressant) | 4.42 | 2.50 – 9.01 |
| Psytx w pt 45 minutes | 6.06 | 3.62 – 10.28 |
| Trazodone (antidepressant) | 12.63 | 6.12 – 31.04 |
| Crisis intervention mental h | 4.71 | 3.02 – 7.72 |
| Age | 0.16 | 0.12 – 0.21 |
| Amoxicillin (antibiotic) | 0.14 | 0.08 – 0.22 |
| Dextroamphetamine amphetamine (ADHD) | 0.01 | 0.01 – 0.02 |
| Ferrous sulfate (iron deficiency) | 0.01 | 0.01 – 0.02 |
| Nitrofurantoin mono macro (antibiotic) | 0.07 | 0.03 – 0.40 |
| Nystatin (antifungal) | 0.03 | 0.01 – 0.06 |
| Oxycodone acetaminophen (opioid pain reliver) | 0.04 | 0.01 – 0.98 |

Two types of services were consistently predictive of a PRTF entry for all age groups: psychotherapy and crisis intervention mental health services. For 13- to 15-year olds, billing for drug screens was additionally associated with entering PRTF. Antipsychotic prescriptions (e.g., Abilify, risperidone) and ADHD prescriptions (e.g., clonidine, guanfacine) were strong predictors of PRTF entry among all age groups. Additional risk factors among children 6- to 15-years old included antiepileptic prescriptions (e.g., oxcarbazepine) and among older children (16-21 years old), antidepressant prescriptions (e.g., fluoxetine, trazodone). Claims for antibiotics were most prevalent among non-PRTF children and thus consistently appeared as a ‘protective’ factor in our predictive model.

### 3.3 Readmission Analysis

Within six months after the initial PRTF discharge, only 47 different services were claimed; the most common of which was nasal drainage. Only three children (all of whom subsequently reenter PRTF care) had a claim for a behavioral and mental health service (i.e., mental health crises intervention).

Table 3 shows statistically significant risk factors from a multivariate logistic regression model. Pharmaceutical claims for antipsychotic, ADHA, and antidepressant medications were all associated with elevated risk of PRTF readmission. The only protective factor was the longer length of stay (in days) in the residential treatment program during the initial entry.

**Table 3.** Post-discharge statistically significant factors associated with PRTF reentry.

| Predictor | aOR | 95% Confidence Interval | P-values |
| --- | --- | --- | --- |
| Days in spell | 0.995 | 0.994 – 0.997 |  |
| Abilify (antipsychotic) | 2.291 | 1.629 – 3.220 |  |
| Bupropion (antidepressant) | 2.133 | 1.092 – 4.164 |  |
| Chlorpromazine (antipsychotic) | 1.810 | 1.049 – 3.123 |  |
| Clonidine (ADHD) | 1.489 | 1.138 – 1.950 |  |
| Desmopressin acetate | 1.446 | 1.012 – 2.066 |  |
| Divalproex sodium (antiepileptic) | 1.451 | 1.021 – 2.063 |  |
| Ibuprofen | 1.708 | 1.152 – 2.534 |  |
| Intuniv (ADHD) | 1.786 | 1.135 – 2.810 |  |
| Mapap (analgesic) | 3.905 | 1.885 – 8.092 |  |
| Mupirocin (antipsychotic) | 1.803 | 1.036 – 3.137 |  |
| Oxcarbazepine (anticonvulsant) | 1.448 | 1.020 – 2.056 |  |
| Quetiapine fumarate (antidepressant) | 1.554 | 1.172 – 2.062 |  |
| Risperidone (antipsychotic) | 1.543 | 1.196 – 1.992 |  |
| Seroquel (antipsychotic) | 2.869 | 1.640 – 5.020 |  |
| Trazodone (antidepressant) | 1.338 | 1.057 - 1.695 |  |

## 4. DISCUSSION

In the present study of Kentucky child and youth Medicaid beneficiaries aged 6-21 we found that placements in PRTFs are relatively rare events, with afewer than 0.5% rate of occurrence. In comparison, a study investigating the rate of PRTF admissions among children between 5 and 17 years of age with a history of maltreatment that were Medicaid beneficiaries in North Carolina reported 1.6% PRTF admission rate.^8^ Another study of Medicaid enrolled children 3-17 years old in Kansas reported 2.5% admission rate.^7^ A study that investigated state variation in the use of out-of-home mental health services among children and youth under age 22 enrolled in Medicaid during 2003 reported 3.9% (North Carolina), 4.2% (Oklahoma), and 6.6% (New Jersey) PRTF admission rates. Overall, our findings suggest that the rate of PRTFs admissions in Kentucky is low, although comparative data are somewhat lacking.

Despite a very low prevalence of PRTF admission, we developed and tested a predictive modelling approach based on an ensemble machine learning method that correctly classified over 90% of children and youth who would enter into a residential care (i.e., true positive) and those who would not (i.e., true negative). To the best of our knowledge, this is the first predictive model that uses Medicaid administrative data with strong performance for predicting beneficiaries’ risk of a PRTF entry. Our ensemble approach to predictive modeling achieved higher accuracy then median accuracy of base learners, confirming its superior predictive value. Nonetheless, more testing as new data become available is needed to further evaluate predictive accuracy.

With regard to medication utilization, consistent with existing literature,^7,13^ we found that polypharmacy was highly associated with increased risk of PRTF admission. Moreover, post discharge polypharmacy prescribing practices persisted. This positive relationship between polypharmacy and risk of PRTF entry potentially raised prior concerns^14^ and suggests further evaluation of physician prescribing practices, sufficiency and timeliness of outpatient medication appointments, especially in vulnerable groups of younger children. Furthermore, systematic antibiotic use among children and adolescents without PRTF entries indicates limited adherence to evidence-based practice guidelines in antibiotic prescribing in outpatient settings.

Three service types were associated with elevated risk of PRTF entry: psychotherapy, crisis intervention mental health services, and billing for drug screen. Prior reports found that over 92% of PRTF-admitted youth in Kansas received some type of outpatient mental health service prior to admission.^7^ The rate of service utilization in our study is considerably lower and practically diminishes during 6 months post discharge. If one assumes that outpatient service utilization is a proxy of mental health need then it may be possible that factors other than clinical need may lead to a PRTF entry.

The results of this study must be viewed in the context of the study’s limitations. First, the study relied on administrative claims data that despite their richness are not constructed for the purpose of research and have reliability and validity concerns. Second, a cross-sectional design was used to evaluate our first aim (predictors of first PRTF admission). Further examination of patterns and predictors of children’s psychiatric hospitalization would benefit from new longitudinal data, that will also allow one to reexamine predictive validity of our model.

### 4.1 Implications

The findings of the present set of analyses of the utilization of PRTFs in Kentucky among Medicaid recipients have several clear implications. First, it is clear that for many children and youth in Kentucky the pathway into admission to a PRTF involves the receipt of psychiatric care, both outpatient and inpatient. In addition, the use of anti-psychotic and anti-manic medications are much more common among youth who are admitted into these residential facilities. These findings suggest that, despite the fact that admission into a PRTF is a relatively rare event in the entire population, it is a predictable event among youth with identified mental health conditions who are receiving care in the community.

Second, there is a surprising drop off in service utilization in the period following discharge from an index admission into a PRTF. A large proportion of youth do not appear to receive any follow-up care in the first 90 days after discharge. More troubling, those that do receive post-discharge care are actually more likely to be re-admitted than those who do not receive any care.

The present findings have both positive and negative implications for the Commonwealth’s public behavioral health system for children and youth. First, placement in PRTFs are relatively rare. This is an intensive and expensive intervention than almost no youth wishes to experience. There is a growing consensus that residential treatment is a worst-case solution for children and youth, so the rarity of the event in Kentucky can be seen as a positive. Further, it does appear that the pathways into PRTF suggests this intervention is used for young people with significant psychiatric needs. While this finding is very limited due to our reliance on service utilization data, it is supported by a review of files, accomplished in the early stage of this analysis. However, to categorially state that PRTFs are serving a clinically indicated sub-population, it would be necessary to have consistent clinical and functional assessment information available.

The novel analytic strategy used in the present study may have implications for other health services researchers in the Commonwealth. Since, placement in a PRTF is a low base rate event, a straight prediction model is difficult. The use of an iterative modeling approach that over-samples the rare cases but compares them to a thousand randomly generating samples, allows for robust and generalizable modeling of rare events.

## Data Availability

All data for this study is restricted.

## Acknowledgments

This work was supported by the State University Partnership (SUP) research program of the Kentucky Cabinet for Health and Family Services (CHFS), and by the University of Kentucky.

## Notes

### Competing Interest Statement

The authors have declared no competing interest.

### Author Declarations

The University of Kentucky institutional review board approved this study.

